# Development and application of Pandemic Projection Measures (PPM) for forecasting the COVID-19 outbreak

**DOI:** 10.1101/2020.05.30.20118158

**Authors:** Shu-Ren Chang

## Abstract

This study aims to provide an accessible and dynamic estimate method to project the Covid-19 trend and hopefully it will help inform policymakers to allocate the needed medical equipment and supplies for saving more lives. A set of newly developed Pandemic Projection Measures (PPM) had been successfully applied to project daily new cases across countries. During the development, numerous trial and error iterations had been performed and then improved with live data. The procedures and computations for the PPM including Uphill Index (UHI), Downhill Indices (DHI), and Error Band Projection (EBP) estimates were explained and discussed along with graphical projections. The PPM was computed with daily live data for the USA, four U.S. states (Illinois, Massachusetts, New Jersey, and New York), France, Italy, Spain, Germany, and China. The results indicated that with the PPM estimations, the daily projections for the future trend were robust to reflect the most plausibility, since the PPM can be updated frequently. With the most up-to-date predictions, governments should be able to monitor the values of UHI and DHI for making a better decision for “flattening the curve”. Based on the empirical data, policymakers should pay more attentions for the following two scenarios: a) When expecting an apex of the outbreak, the UHI is higher than 1.20; and b) After passing a peak day, the DHI is still larger than 0.925. The applications of the PPM estimates are not designed for a one-time projection rather than updated frequently to improve the prediction precisions. With the same concepts from the PPM computations, the peak day and the number of new deaths could be predictable if more data are collected. Like many country leaders saying, “We will win the battle of coronavirus pandemic”, the author hopes to use this easily applicable estimate method to save more lives and to win. The results, currently presented with the data on April 16 and 17, 2020, were only used to explain how to apply the PPM estimates for predictions. The outdated results should not be used to compared with today’s outbreak trend.

## Introduction

Since December 2019, Coronavirus (Covid-19) has caused 191,061 deaths across 210 countries and territories with a dramatic daily increment up to 2,725,920 cases today [1]. It is believed to have more higher infected cases or even more deaths, as many infected died before they were tested. Many more infected might have not been screened or have only mild or no symptoms at all. In China, the Covid-19 outbreak overwhelmed their medical facilities and staff, and their medical equipment and supplies fell woefully short. The number of the deceased in China was underestimated due to lack of testing, screening, and honest reporting. People over the globe are very anxious about this devastating pandemic and all eager to know when it would end soon. On April 13, 2020, the U.S. President Donald Trump claimed to reopen the U.S. economy by May 1 to help citizens back to normal. However, public health care experts debated over the question of when to reopen portions of the U.S. economy. They speculated that the reopening may be unrealistic by May 1 [2]. Reopening to save the economy has been debated a lot more in many other countries.

Many mathematicians and statisticians proposed different kinds of models available to predict outbreak trends for contagious disease such as severe acute respiratory syndrome (SARS), Middle East respiratory syndrome (MERS), or Covid-19 [3]. Most statistical models developed provide a one-time deal for the predictions for upcoming weeks or months [4] [5]. Some more promising research works have been used to predict total deaths, the needed hospital resource use, the number of beds, intensive care unit (ICU) beds, and invasive ventilators through web-based estimations [4] [6].

Governments from countries across the world are relying on mathematical projections to help guide decisions for early preparation in this pandemic [7]. The death numbers could dramatically roar as the needed medical supplies and lifesaving equipment fall shorts. Many different growth models have been explored to model the pandemic data for projections on the numbers of the infected [8]. However, more dynamic or even daily updated ones are still needed to inform governments for better policy-decisions. Therefore, it is an urgent need to develop estimate methods that help provide a quick turnaround projection to prepare for the worst.

Much evidence supports the cases of Covid-19 are growing exponential and many epidemiologists have suggested using social distancing, quarantine, and isolation to flatten the upcoming curve for slowing down its quick spread [9]. Slowing down the spread can also avoid overwhelming health care professionals and will allow them to have more time to allocate more medical supplies to save more lives. An interesting research discussed how an exponential function or logistics growth model could be used to model contagious disease like Ebola virus [10]. Some research believed that many pandemics could spread exponentially and the trend of Covid-19 outbreak could be fitted in exponential growth models [10] [11] [12] [13]. Some statisticians discussed the estimates methods using exponential grow and found that it was promising for projecting pandemics like SARS and Covid-19 [11] [12]. Many of the statistics models for projections referred by news media did not provide much detailed explanations with mathematical computations behind online instant resources. Most of them provided a one-time prediction for a few weeks or months, which might not accurately reflect the most recent reality. There is an urgent need for policymakers who reply on much simpler accessible prediction models with visual presentations to help people quickly understand the pandemic trend of Covid-19.

This purpose of this study is three-fold: (a) to provide a fast and dynamic projection method to forecast Covid-19 outbreak trends; (b) to provide a user-friendly method that is easily replicable and accessible for their own regions, cities, countries, etc.; and (c) to provide for an instant update with data visualization solutions to help predict the trends in their own regions.

### Data sources and analysis tools

Two data sets were downloaded from online resources. Both websites updated daily are free and open for the public to download. Data contained the number of new cases, total cases, new deaths, and total deaths from all countries around the world sponsored by Global Change Data Lab [14]. Data from the U.S. States are available for download from https://github.com/ sponsored by The New York Times [15]. The SAS® Enterprise Guide® program was used to read in the data from websites and then the Microsoft Excel program was used to compute the Pandemic Projection Measures (PPM). The SAS® SGPLOT procedure, part of Output Delivery System (ODS) Statistical Graphics then was used to plot the PPM estimates with Needle Plots to better present current new cases, projected new cases, and new deaths for countries and states. Many data resources are available for download. However, there is no one central office taking care of the reporting consistence for Covid-19 cases all regions. There might be some discrepancy for some countries/ regions for the daily reporting.

### Research methods

Many studies with much evidence suggested that an exponential growth pattern can be used to model the cases of infectious respiratory diseases such as SARS and Covid-19 [12] [16]. Therefore, in this study, it is assumed that the simulated Covid-19 new cases would follow an exponential grow trend before the peak day and then decline exponentially after the peak day for a region. Based on the assumption for exponential grow and decay, a set of pandemic projection measures (PPM) were developed to project daily new cases.

### Development procedures: Numerous trial and error iterations

During the development process, a set of live data from USA, four U.S. states (Illinois, Massachusetts, New Jersey, and New York), France, Italy, Spain, Germany, and China had been applied and tried out with the SAS to model data and plot the outbreak trends. Numerous trial and error iterations using the PPM on real data from different geographic regions have been executed to simulate daily new cases. The simulated cases then were plotted using the SAS to predict plausible outbreak trends. Numerous modification iterations on the PPM estimates along the error band boundaries have been performed to improve the plausibility. Finally, graphical presentations using the SAS SGPLOT procedure were generated to present the improved PPM estimates. The first country for the simulation is China, which data provided the best sample for data modeling, because China has almost gone through a complete devastating outbreak cycle.

The PPM contained three measures: Uphill Index (UHI), Downhill Index (DHI), Error Band Projection (EBP). Before the PPM formulas are presented, growth factor, growth decay, and exponential growth/decay are explained first.

### Exponential growth and decay formulae

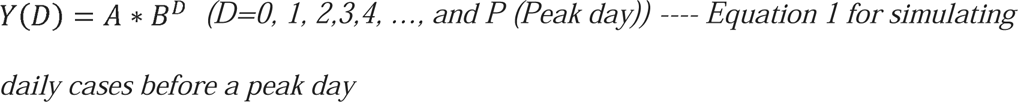

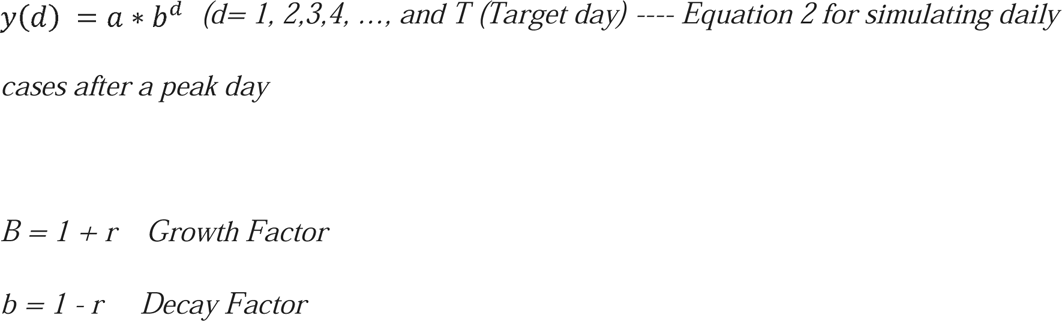

> D is the number of outbreak days before the peak day in the region;

> d is the number of outbreak days after the peak day in the region;

> *Y*(D) or y(d) is the value (i.e., the number of new cases) at the total D or d day;

> *A* is the initial value at time D = 0 (Initial amount before measuring growth or decay);

> *a =* Y(D) is the maximum simulated value. The a should be replaced by Y(D) to simulate daily new cases after the peak day; and

> r is the growth rate (when r > 0) or decay rate (when r < 0) (most often represented as a percentage and expressed as a decimal) [17].

In this study, *Y*(D) or y(d) is the target number of the simulated daily new cases before and after the peak day. In the function above, the D or d represents the number of days the growth or decay factor is multiplied. The a represents the starting value such as the starting population being confirmed by Covid-19. The b represents the growth/decay factor. If b > 1, the function represents exponential growth. If 0 < b < 1, the function represents exponential decay. The r represents the growth or decay factor with a decimal or percent. Before the peak day of the outbreak, a grow factor can be computed for every day. Likewise, a decay factor can be calculated after the peak day. A growth of 21% is a growth factor of 1 + 0.21 = 1.21. A decay of 19% is a decay factor of 1 - 0.19 = 0. 81.

### Application of growth and decay factors to daily new cases

In the definition above, the growth/decay factor is the factor by which a quantity multiplies itself over time. However, in this study, each day a grow or decay factor is computed as “every day’s new cases / (divided by) new cases on the previous day”. For example, if the measure growing by 8% for a day has a growth factor of 1.08, and vice versa, if the measure deceasing by 15% for a day has a decay factor of .85. A growth factor greater than 1 indicates an increase, whereas one that ranges between 0 and 1 indicates a sign of deceasing. A growth factor constantly above 1 could signal exponential growth. However, in some cases, it might decease in one day and increase dramatically on the next. That yields a substantial grow factor like 2, 3, 4, 5, … which is usually considered as an outlier eligible to be removed from calculations to avoid an over-inflated estimate. Whether including or excluding each daily growth/decay factor that looks like an outlier or not could be a big challenge as at the earlier stage of the outbreak the case numbers went up and down dramatically sometimes. It might inflate the simulation estimations dramatically. Thus, some of the first 10 grow factors might be excluded for the computations for simulations.

### Pandemic Projection Measure (PPM)

The three newly developed PPM including Uphill Index (UHI), Downhill Index (DHI), and Error Band Projection (EBP) are explained below.

Before computing the following PPM, the first step is to identify a peak day with the highest number among multiple daily new cases. Once the peak day is identified, each grow factor before the peak day can be computed, and then all grow factors can be averaged. That is, the Uphill index (UHI), the average of all daily growth factors excluding outliers with values larger than 2 or 3. Its formula is presented below:

Uphill Index (UHI) 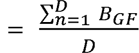

> D is the upper limit as the total number of outbreak days before the peak day; n is the lower limit starting from Day one before the peak day; and

> B_GF_ is the growth factor before the peak day

The Downhill Index (DHI) is the average of all daily decay factors from the peak day to the most current target day. Any value larger than 2 or 3 might be considered as an outlier eligible for removal as the number went up or down dramatically on any adjacent day. The DHI can be expressed below:

Downhill Index (UHI) 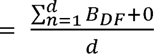 (A pseudo-value, 0, is used to substitute the unknow DHI in the future)

> d is the upper limit as the total number of outbreak days after the peak day;

> n is the lower limit starting from Day one after the peak day to the most current; and

> B_DF_ is the decay factor after the peak day

Each of the simulated daily new cases can be produced, once both UHI and DHI for a region are computed from the live data. In Equation 1, the B can be replaced with the UHI to generate a simulated case for each day before the peak day. In Equation 2, the b can be replaced with the DHI to get a simulated one for each day after the peak day. With a few iterations, a staring value with a decimal or integer can be determined, after the simulated maximum new cases matched closely to the maximum number of daily new cases on the peak day that is identified from live data from the region. If there are only less than 5 days of DHIs available after the peak day, another or more pseudo values of 1 might be needed for the simulation. Figure 1 presented how the results of the UHI, DHI, and EBP using China’s live data were applied in the simulation.

**Figure 1:**
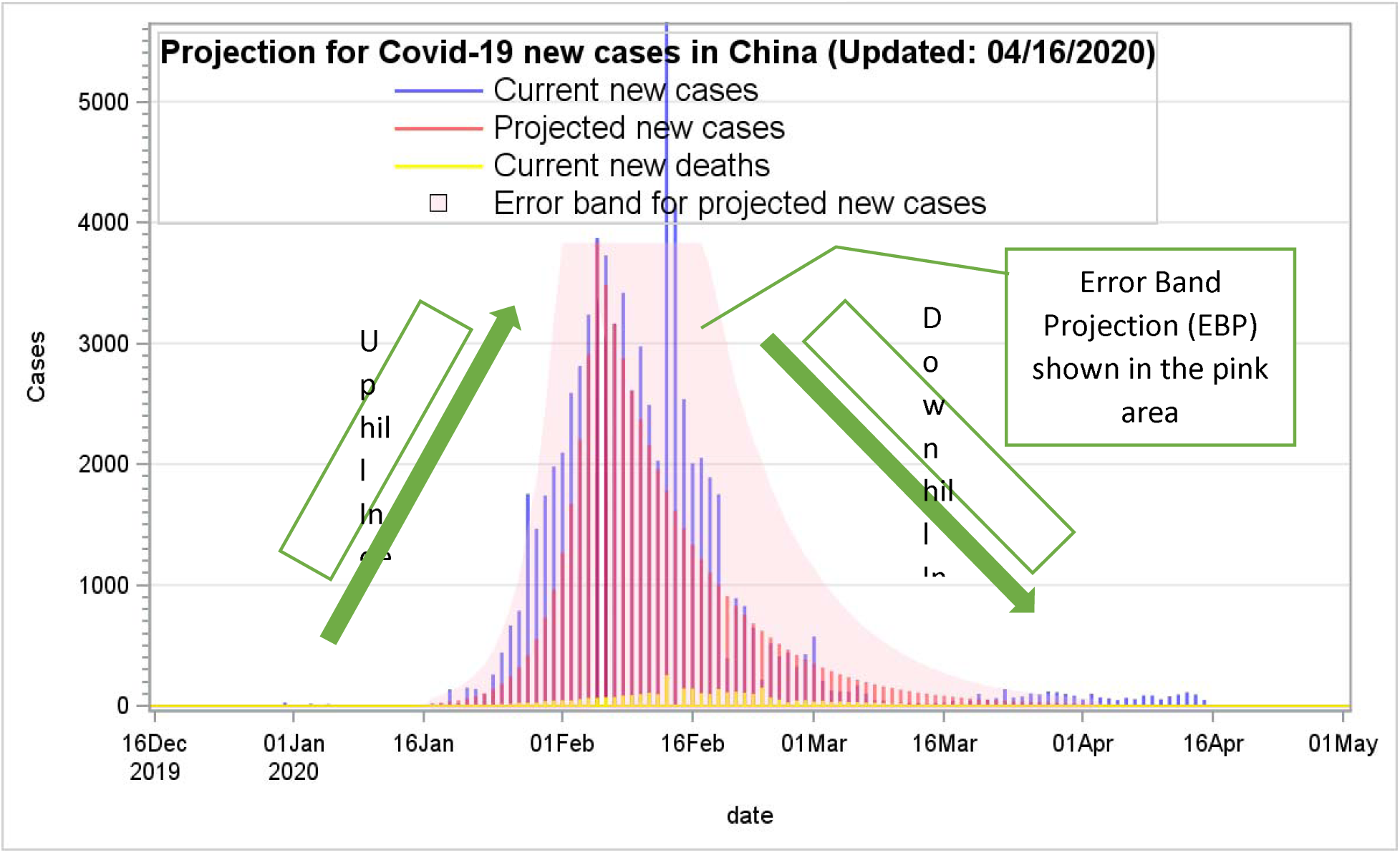
Projection for Covid-19 new cases in China.

### Error Band Estimates

Every measurement or estimation has errors. Measurement error is quantified to estimate the tolerable errors to improve the precision in many areas like physics, economics, biometrics, psychometrics, mathematics, psychology, education, testing, measurement, etc. Measurement error and its error band computation are commonly applied by many professionals with some computational variations for their specific needs to inform their professionals in clinics, teaching, treatments, interventions, or experiments, etc. to improve their practice. In education, Standard Error of Measurement (SEM) with confidence intervals (CI), equivalent to error band, is widely used [18], while error measurement with confident intervals is commonly used for sampling error estimates in labor statistics [19]. Error measurement, measurement error, SEM, bias, imprecision, relative error, total error, total analytic error, etc. are all with different names for errors, but their computations and concepts are somehow similar [20]. Its simple formula in this study can be expressed as: Error (Bias) % = (Average absolute deviation from the simulated value/ (divided by) targeted simulated value) × 100 [21]. In this study, the Error Band Projection (EBP) is adapted from the Error (Bias) and the Mean Absolute Percentage Error (MAPE) [22].

Then the upper and lower bounds of the Error band for the simulated cases can be computed using the formula below:

### Error Band Projection (EBP) formula

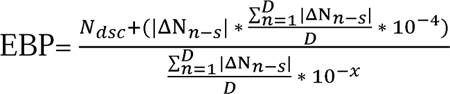 (x ≥ 3, x = 3 when new cases of the peak day are less than 3×10^4^ or x =4 when the ones are greater than 3×10^4^)

> N_dsc_ is the number of daily simulated new cases;

> ΔN_n-s_ is the number difference between new Covid-19 cases and simulated cases;

> x is an adjustable value that can be determined by a user for how wide an error band will expect to project the upcoming new cases. However, an increment of one for x is recommended; and

> D is the upper limit as the total number of outbreak days before the peak day

Upper Bound EBP (UEBP) 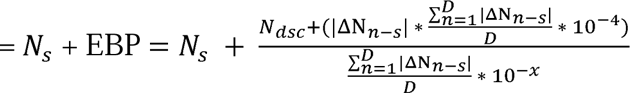 (UEBP > 0),

N_s_ = the number of simulated Covid-19 cases;

Lower Bound EBP (LEBP) 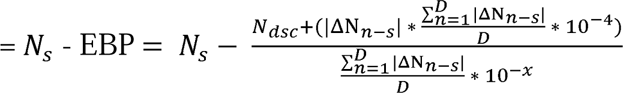

(LEBP ≥ 0)

The EBP is the product of the average absolute deviation from the simulated numbers and the average absolute deviation from the simulated cases divided by the number of the targeted. Therefore, the value of EBP is designed to be slightly inflated to allow more predictability. In other words, the estimates of the error band are expected to be rather larger to cover more unexpended new cases. In some occasions, as days go by, a new peak day might need to be re-define and all PPM estimates will need to be re-calculated for a better precision as more new cases may soar in the following days.

## Results

This study is to present the newly developed pandemic projection methods to simulate when the convid-19 outbreak might be faded out. With live data, a set of pandemic projection measures (PPM) had been developed and successfully applied to the USA, four U.S. states (Illinois, Massachusetts, New Jersey, and New York), France, Italy, Spain, Germany, and China. The results with UHI and DHI were presented and discussed for the six countries and four U.S. states. However, the graphic presentations along with the PPE were further discussed only for USA, Illinois in U.S., France, Italy, and Spain. The results below were used to explain how PPM were computed and applicable to predict the outbreak trend. However, the projection results on April 16 and 17 were outdated now. The PPM should be updated to reflect the most current reality.

### The UHI and DHI: indicators for the rate of exponential growth/decay

From the empirically data in this study, the UHI and DHI are robust indicators for the rates of the outbreak. As shown in Table 1, the higher UHI, the dramatical growth will be like New Jersey. On April 17, its UHI was 1.397, indicating that the average growth of all the days before the peak day was quite dramatically, since most of the UHI ranged from 1.2 to 1.3. The lower UHI means a lower growth in average for all the days before the peak. The UHI and DHI can be updated frequently per day. Regarding the DHI, the higher DHI, the longer the outbreak will linger, as the higher DHI indicated that their new cases are still not fading out yet. On April 17, Spain had an estimated DHI value of 0.928, while Italy had the one of 0.944. These two higher DHI values indicated at the time the outbreak might extend a bit longer period as most of DHI values range from 0.8 to 0.9. The largest UHI and DHI was 1.394 and 0.944 respectively. Based on the empirical data, governments might alert their citizens in the following two scenarios: a) Before the apex of the outbreak, the UHI is higher than 1.20, which indicates that the current exponential grow might overwhelm their health care system soon; and b) After the peak day, the DHI is still larger than 0.925, which indicates that the outbreak might not fade away soon. In these two scenarios, more restrict policy might need to be imposed, such as social distancing, self-quarantine, wearing face masks, etc.

**Table 1:**
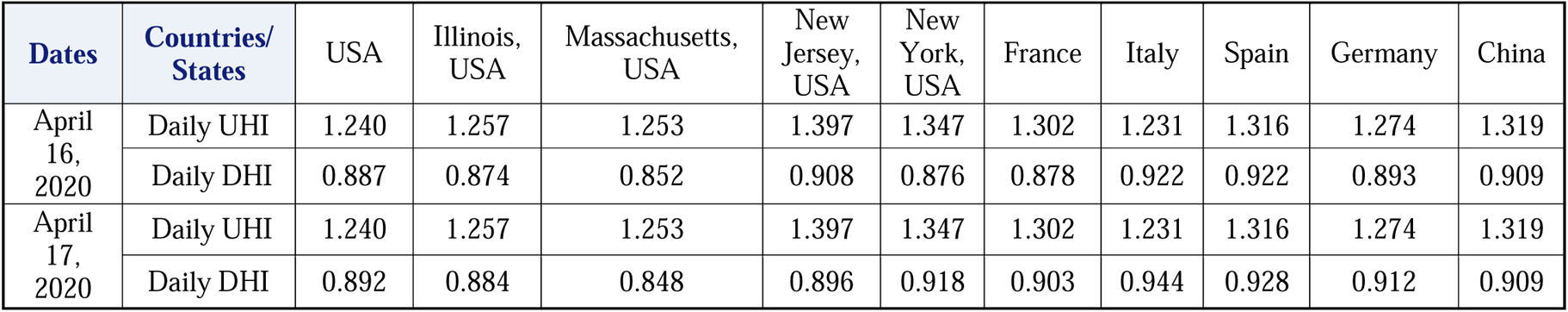
The Daily UHI and DHI across counties and U.S. states on April 16 and 17, 2020.

### United States

On April 13, 2020, U.S. President, Donald Trump hoped to reopen the U.S. economy by May 1. Will it be possible to see the Covid-19 fade away by then? Based on the results from the PPM analyses, the answer is: Not so fast. Let us assume the peak day was April 11 with new cases of 35,527. As shown in Figure 2 and Table 1, if after the peak day, it begins to decline exponentially with a DHI value of 0.892. On the day of May 1, there will be still a minimum of 3,731 new cases. If we look at the error band (shaded in pink), there will be possibly up to 13,143 cases on May 1. Using the PPM estimations, it can help provide a concrete projection with the visual presentation to inform policymakers to make a realistic decision. However, the USA has a total population of 330 million and has more than big10 cities exceeding one million in population. Some cities’ Covid-19 cases are growing exponentially; others’ ones are decaying. It is also very challenged to make a fair prediction using one visual graphic presentation for the U.S. with such a big population and so many big cities together.

**Figure 2:**
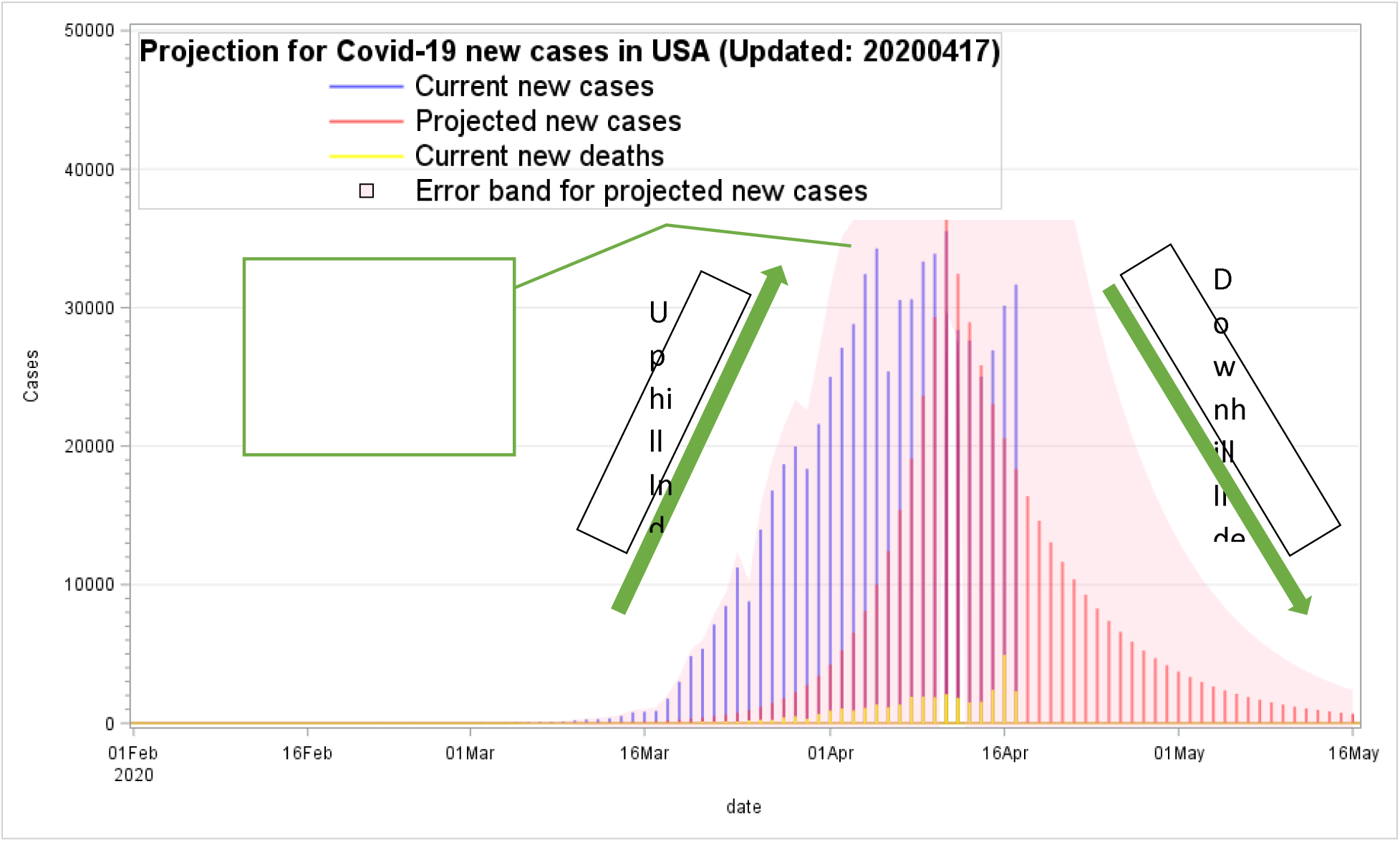
Projection for Covid-19 new cases in the United States.

### Spain

On January 31, 2020, the Covid-19 pandemic was confirmed to have spread to Spain, after a German tourist was tested positive for SARS-CoV-2 in La Gomera, Canary Islands. As presented in Figures 3.1 and 3.2 for Spain, at the end of February, the case numbers were beginning to increase dramatically, and it seemed to follow along the exponential growth trend before the peak day in April 2020. After the peak day (as being set at this moment), the numbers of daily new cases were decreasing exponentially along with the simulated trend, and all fell in the error bands. On April 15, there were 3,045 new cases with a DHI value of 0.872. As shown in Figure 3.2, Tables 2.1 and 2.2, two more higher case numbers with 5,092 and 5,183 were observed on April 16 and 17. The DHIs were accordingly increased from 0.872 (April 15) to 0.922 (on April 16) and then to 0.928 (on April 17). After new data were included, the upper error band also expanded much higher accordingly to provide a better prediction for the upcoming. The results indicated the PPM estimates were very adaptive and robust for predictability when more higher number cases rose. Based on the data collected on April 16, the predicted outbreak decline trend might likely occur around May 16. However, after a much higher new case number on April 17 was added, with the PPM estimates, the predicted outbreak will not be expected to be ending soon and might extend a bit longer after Mat 16 and beyond.

**Figure 3.1:**
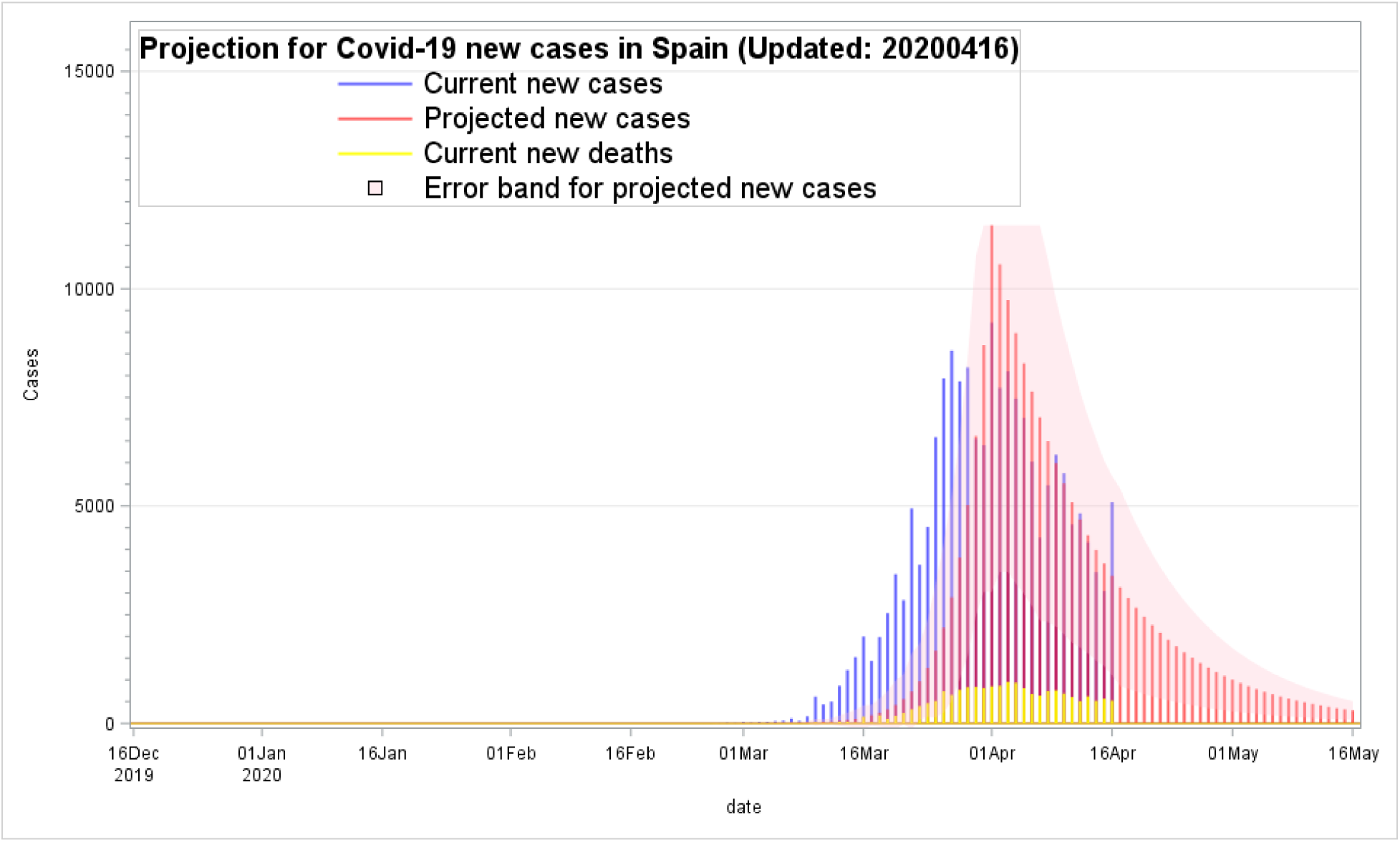
Projection for Covid-19 new cases in Spain.

**Figure 3.2:**
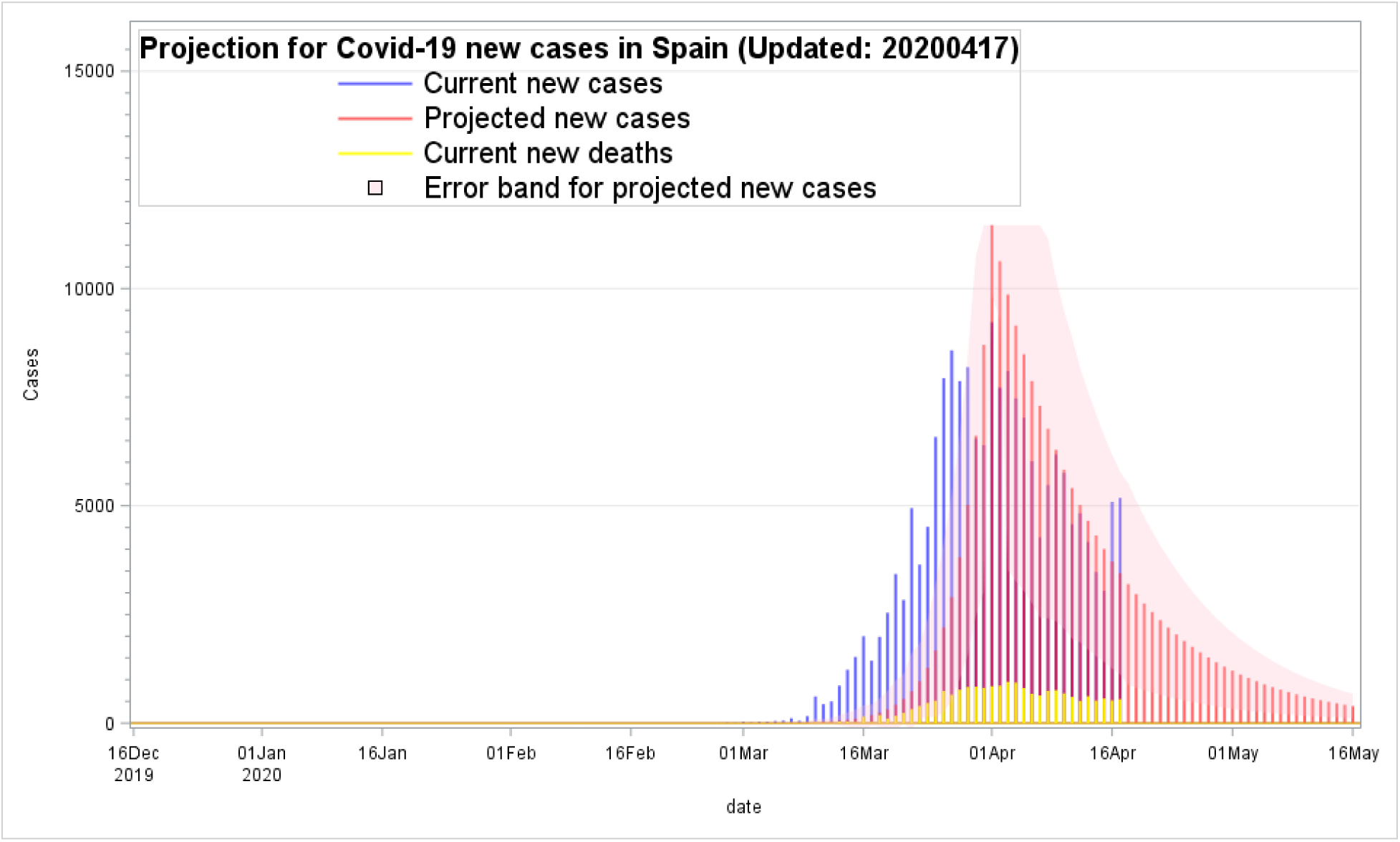
Projection for Covid-19 new cases in Spain.

**Table 2.1:** The Pandemic Projection Measures (PPM) estimations on April 16, 2020 for Spain.

| Date | Daily New cases | PPM estimations on April 16, 2020 |  |  |  |  |
| --- | --- | --- | --- | --- | --- | --- |
|  |  | Simulated cases | Error Band | Upper Error Band | Lower Error Band | UHI/DHI |
| 2/28/2020 | 13.0 | 1.3 | 2.0 | 3.3 | 0.0 | 1.316 |
| 2/29/2020 | 9.0 | 1.7 | 1.8 | 3.5 | 0.0 | 1.316 |
| 3/1/2020 | 32.0 | 2.3 | 4.4 | 6.7 | 0.0 | 1.316 |
| 3/2/2020 | 17.0 | 3.0 | 3.3 | 6.3 | 0.0 | 1.316 |
| 3/3/2020 | 31.0 | 4.0 | 5.2 | 9.1 | 0.0 | 1.316 |
| 3/4/2020 | 37.0 | 5.2 | 6.4 | 11.6 | 0.0 | 1.316 |
| 3/5/2020 | 49.0 | 6.8 | 8.5 | 15.3 | 0.0 | 1.316 |
| 3/6/2020 | 61.0 | 9.0 | 10.8 | 19.9 | 0.0 | 1.316 |
| 3/7/2020 | 113.0 | 11.9 | 17.5 | 29.4 | 0.0 | 1.316 |
| 3/8/2020 | 56.0 | 15.6 | 13.8 | 29.4 | 0.0 | 1.316 |
| 3/9/2020 | 159.0 | 20.6 | 26.7 | 47.3 | 0.0 | 1.316 |
| Intentionally omitted due to a limited space |  |  |  |  |  |  |
| 3/30/2020 | 6549.0 | 6611.1 | 4141.9 | 10753.0 | 2469.2 | 1.316 |
| 3/31/2020 | 6398.0 | 8702.8 | 5674.6 | 11456.3 | 3028.2 | 1.316 |
| 4/1/2020 | 9222.0 | 11456.3 | 5674.6 | 11456.3 | 3028.2 | 1.316 |
| 4/2/2020 | 7719.0 | 10563.5 | 6257.0 | 11456.3 | 3483.3 | 0.922 |
| 4/3/2020 | 8102.0 | 9740.4 | 6257.0 | 11456.3 | 3483.3 | 0.922 |
| 4/4/2020 | 7472.0 | 8981.3 | 5769.3 | 11456.3 | 3212.0 | 0.922 |
| 4/5/2020 | 7026.0 | 8281.5 | 5306.1 | 11456.3 | 2975.3 | 0.922 |
| 4/6/2020 | 6023.0 | 7636.1 | 4938.2 | 11456.3 | 2697.9 | 0.922 |
| 4/7/2020 | 4273.0 | 7041.1 | 4681.5 | 11456.3 | 2359.6 | 0.922 |
| 4/8/2020 | 5478.0 | 6492.4 | 4162.9 | 10655.3 | 2329.6 | 0.922 |
| 4/9/2020 | 6180.0 | 5986.5 | 3764.3 | 9750.8 | 2222.2 | 0.922 |
| 4/10/2020 | 5756.0 | 5520.0 | 3476.7 | 8996.7 | 2043.3 | 0.922 |
| 4/11/2020 | 4576.0 | 5089.9 | 3235.4 | 8325.3 | 1854.4 | 0.922 |
| 4/12/2020 | 4830.0 | 4693.2 | 2949.6 | 7642.8 | 1743.6 | 0.922 |
| 4/13/2020 | 4167.0 | 4327.5 | 2723.2 | 7050.7 | 1604.3 | 0.922 |
| 4/14/2020 | 3477.0 | 3990.3 | 2547.5 | 6537.8 | 1442.8 | 0.922 |
| 4/15/2020 | 3045.0 | 3679.3 | 2365.1 | 6044.4 | 1314.3 | 0.922 |
| 4/16/2020 | 5092.0 | 3392.6 | 2292.2 | 5684.9 | 1100.4 | 0.922 |
| 4/17/2020 |  | 3128.3 | 2269.8 | 5398.0 | 858.5 | 0.922 |
| 4/18/2020 |  | 2884.5 | 2092.9 | 4977.4 | 791.6 | 0.922 |
| Intentionally omitted due to a limited space |  |  |  |  |  |  |
| 5/16/2020 |  | 297.5 | 215.9 | 513.4 | 81.7 | 0.922 |

**Table 2.2:** The Pandemic Projection Measures (PPM) estimations on April 17, 2020 for Spain.

| Date | Daily New cases | PPM estimations on April 17, 2020 |  |  |  |  |
| --- | --- | --- | --- | --- | --- | --- |
|  |  | Simulated cases | Error Band | Upper Error Band | Lower Error Band | UHI/DHI |
| 2/28/2020 | 13.0 | 1.3 | 2.0 | 3.3 | 0.0 | 1.316 |
| 2/29/2020 | 9.0 | 1.7 | 1.8 | 3.5 | 0.0 | 1.316 |
| 3/1/2020 | 32.0 | 2.3 | 4.4 | 6.7 | 0.0 | 1.316 |
| 3/2/2020 | 17.0 | 3.0 | 3.3 | 6.3 | 0.0 | 1.316 |
| 3/3/2020 | 31.0 | 4.0 | 5.2 | 9.1 | 0.0 | 1.316 |
| 3/4/2020 | 37.0 | 5.2 | 6.4 | 11.6 | 0.0 | 1.316 |
| 3/5/2020 | 49.0 | 6.8 | 8.5 | 15.3 | 0.0 | 1.316 |
| 3/6/2020 | 61.0 | 9.0 | 10.8 | 19.9 | 0.0 | 1.316 |
| 3/7/2020 | 113.0 | 11.9 | 17.5 | 29.4 | 0.0 | 1.316 |
| 3/8/2020 | 56.0 | 15.6 | 13.8 | 29.4 | 0.0 | 1.316 |
| 3/9/2020 | 159.0 | 20.6 | 26.7 | 47.3 | 0.0 | 1.316 |
| Intentionally omitted due to a limited space |  |  |  |  |  |  |
| 3/30/2020 | 6549.0 | 6611.1 | 4141.9 | 10753.0 | 2469.2 | 1.316 |
| 3/31/2020 | 6398.0 | 8702.8 | 5674.6 | 11456.3 | 3028.2 | 1.316 |
| 4/1/2020 | 9222.0 | 11456.3 | 5674.6 | 11456.3 | 3028.2 | 1.316 |
| 4/2/2020 | 7719.0 | 10628.1 | 6343.7 | 11456.3 | 3516.1 | 0.928 |
| 4/3/2020 | 8102.0 | 9859.8 | 6343.7 | 11456.3 | 3516.1 | 0.928 |
| 4/4/2020 | 7472.0 | 9147.0 | 5889.5 | 11456.3 | 3257.5 | 0.928 |
| 4/5/2020 | 7026.0 | 8485.8 | 5454.4 | 11456.3 | 3031.4 | 0.928 |
| 4/6/2020 | 6023.0 | 7872.3 | 5109.6 | 11456.3 | 2762.8 | 0.928 |
| 4/7/2020 | 4273.0 | 7303.2 | 4871.7 | 11456.3 | 2431.6 | 0.928 |
| 4/8/2020 | 5478.0 | 6775.3 | 4368.1 | 11143.4 | 2407.2 | 0.928 |
| 4/9/2020 | 6180.0 | 6285.5 | 3942.5 | 10228.0 | 2343.0 | 0.928 |
| 4/10/2020 | 5756.0 | 5831.1 | 3655.2 | 9486.4 | 2175.9 | 0.928 |
| 4/11/2020 | 4576.0 | 5409.6 | 3467.4 | 8877.0 | 1942.2 | 0.928 |
| 4/12/2020 | 4830.0 | 5018.5 | 3158.3 | 8176.8 | 1860.3 | 0.928 |
| 4/13/2020 | 4167.0 | 4655.7 | 2961.3 | 7617.1 | 1694.4 | 0.928 |
| 4/14/2020 | 3477.0 | 4319.2 | 2786.1 | 7105.3 | 1533.0 | 0.928 |
| 4/15/2020 | 3045.0 | 4006.9 | 2602.8 | 6609.7 | 1404.2 | 0.928 |
| 4/16/2020 | 5092.0 | 3717.3 | 2462.9 | 6180.1 | 1254.4 | 0.928 |
| 4/17/2020 | **5183.0** | 3448.6 | 2330.7 | 5779.3 | 1117.8 | 0.928 |
| 4/18/2020 |  | 3199.3 | 2321.3 | 5520.5 | 878.0 | 0.928 |
| Intentionally omitted due to a limited space |  |  |  |  |  |  |
| 5/16/2020 |  | 391.4 | 284.0 | 675.3 | 107.4 | 0.928 |

### Illinois in the United States

As shown in Figures 4.1 and 4.2, based on the PPM estimates using the mostly current data, the results were quite robust in the prediction for the outbreak trend after April 17. The error band estimates between upper and lower bounds well predicted the unexpectedly higher new case numbers like the suddenly increasing ones on April 17. Furthermore, the area of error bands expands much wider after new data from April 17 was included. The number of new cases increased was quite dramatically from 1,141 on April 16 to 1,841 on April 17. The DHI values also increased from 0.874 on April 16 to 0.884 on April 17. This PPM’s prediction with the dynamics DHI was quite sensitive and adaptive after new data was included. The Downhill Index (DHI) should be updated daily to include new information for better plausibility. Unlike other prediction models, most of them only provided a one-time projection for the upcoming weeks or months. In Illinois, a new peak day can be redefined and modified from the current one from April 8 to 17, as this new peak day could provide a better prediction. However, if the peak day was reset, all the UHI, DHI, and error bands will also need to be re-computed for an up-to-date prediction.

**Figure 4.1:**
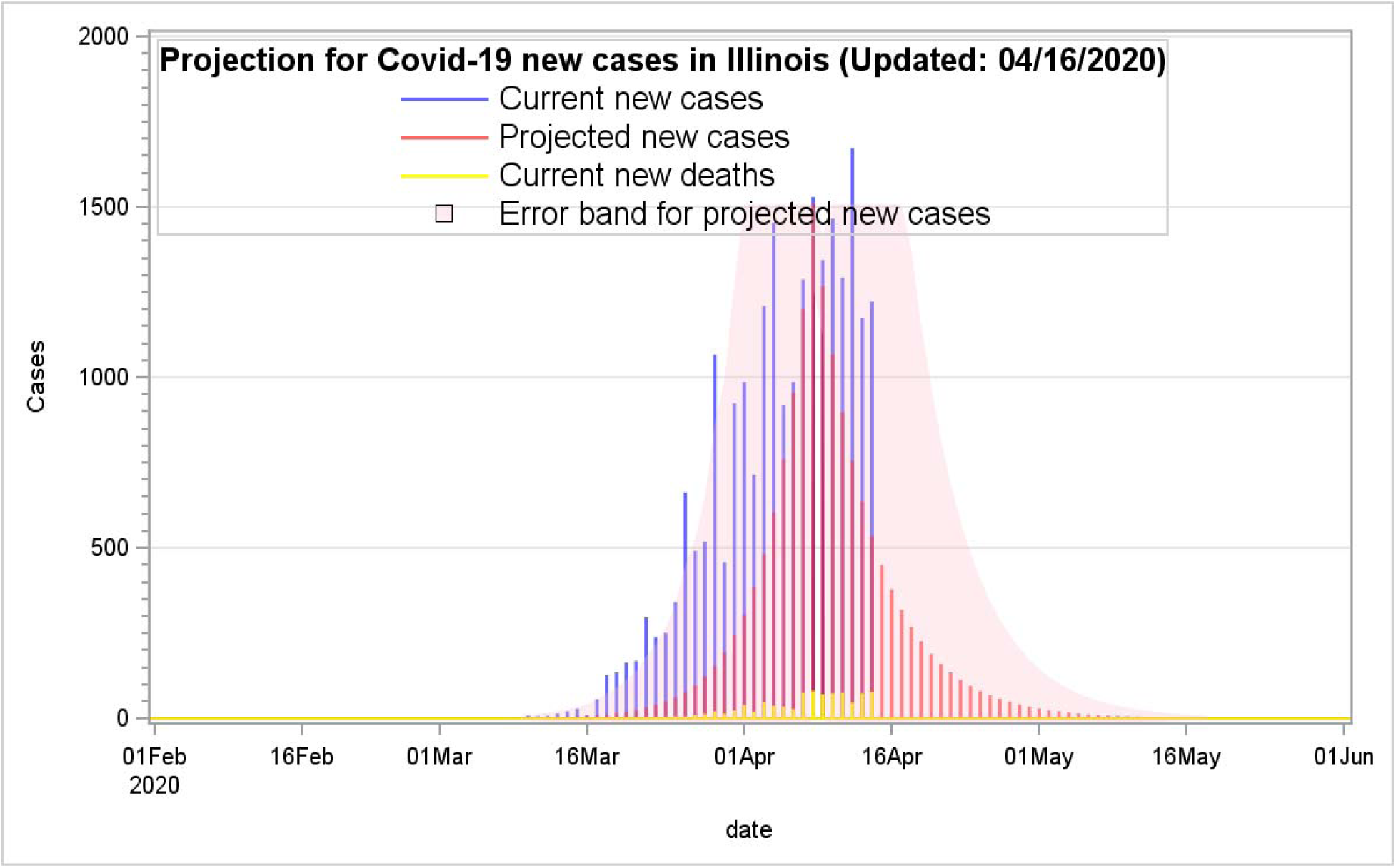
Projection for Covid-19 new cases in Illinois, the United States.

**Figure 4.2:**
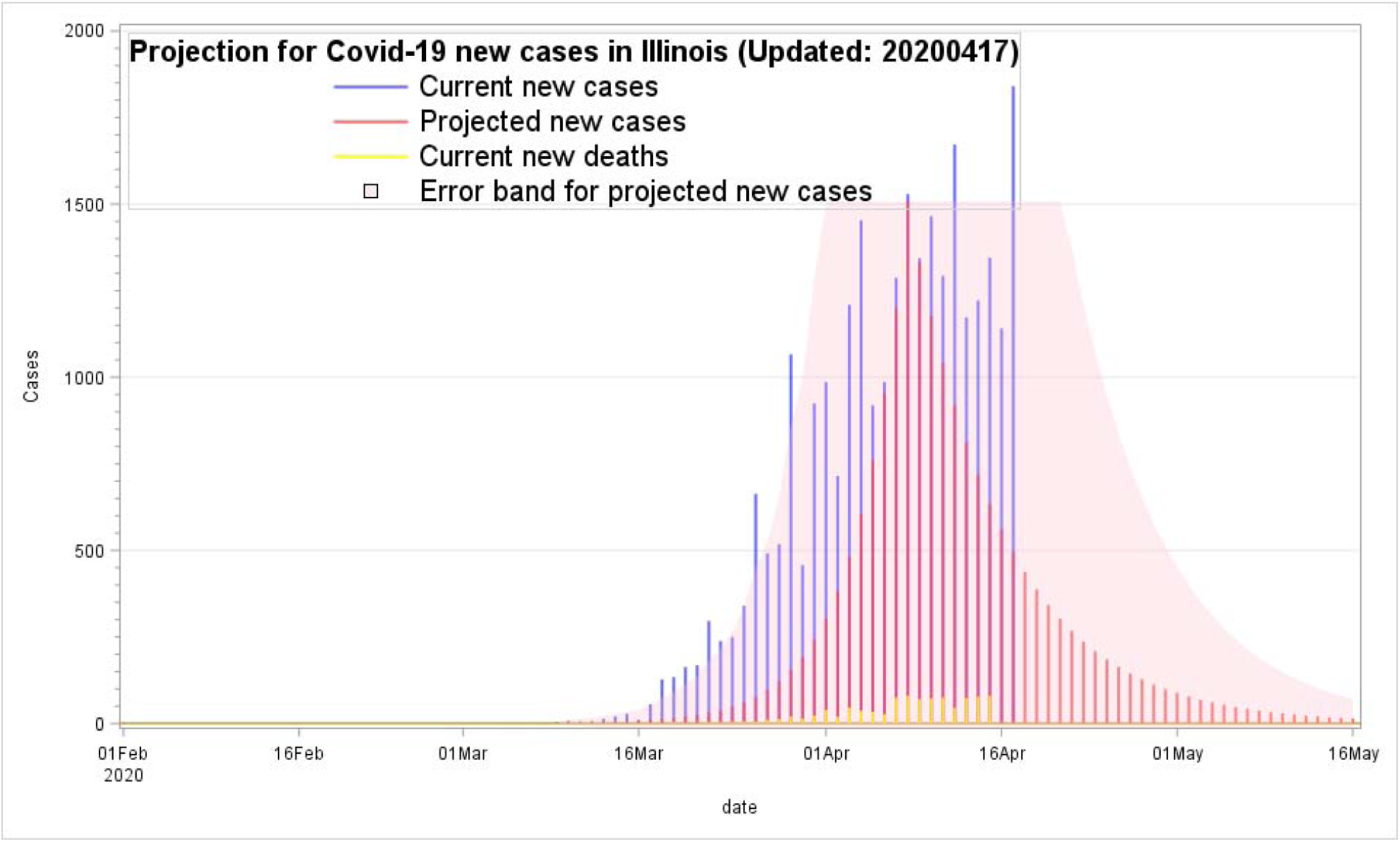
Projection for Covid-19 new cases in Illinois, the United States.

### Italy

With a lingering trend of the covid-19 outbreak in Italy during the end of March as shown in Figure 5.1, the peak day was reset from March 22 to March 27 for a better prediction precision for the outbreak trend. The error band was expanded as a new daily higher case number of 3,786 was added on April 17. The new estimation for a new trend indicated the Covid-19 outbreak might still not fade out quickly until June as shown one in Figure 5.2. The DHI was increased from 0.921 on April 16 to 0.944 on April 17. The DHI value, 0.944, estimated on April 17 was higher than the one, 0.921 estimated on April 16. In other words, based on the newly updated DHI with a higher value on April 17, the higher DHI suggested that the Covid-19 outbreak could last a bit longer. The error band on April 17 will accordingly expend a bit higher than the one on April 16. These DHIs, error band, and graphical presentations were synchronizing to tell a compelling story, but might not be good news for what will likely happen in the next few weeks.

**Figure 5.1:**
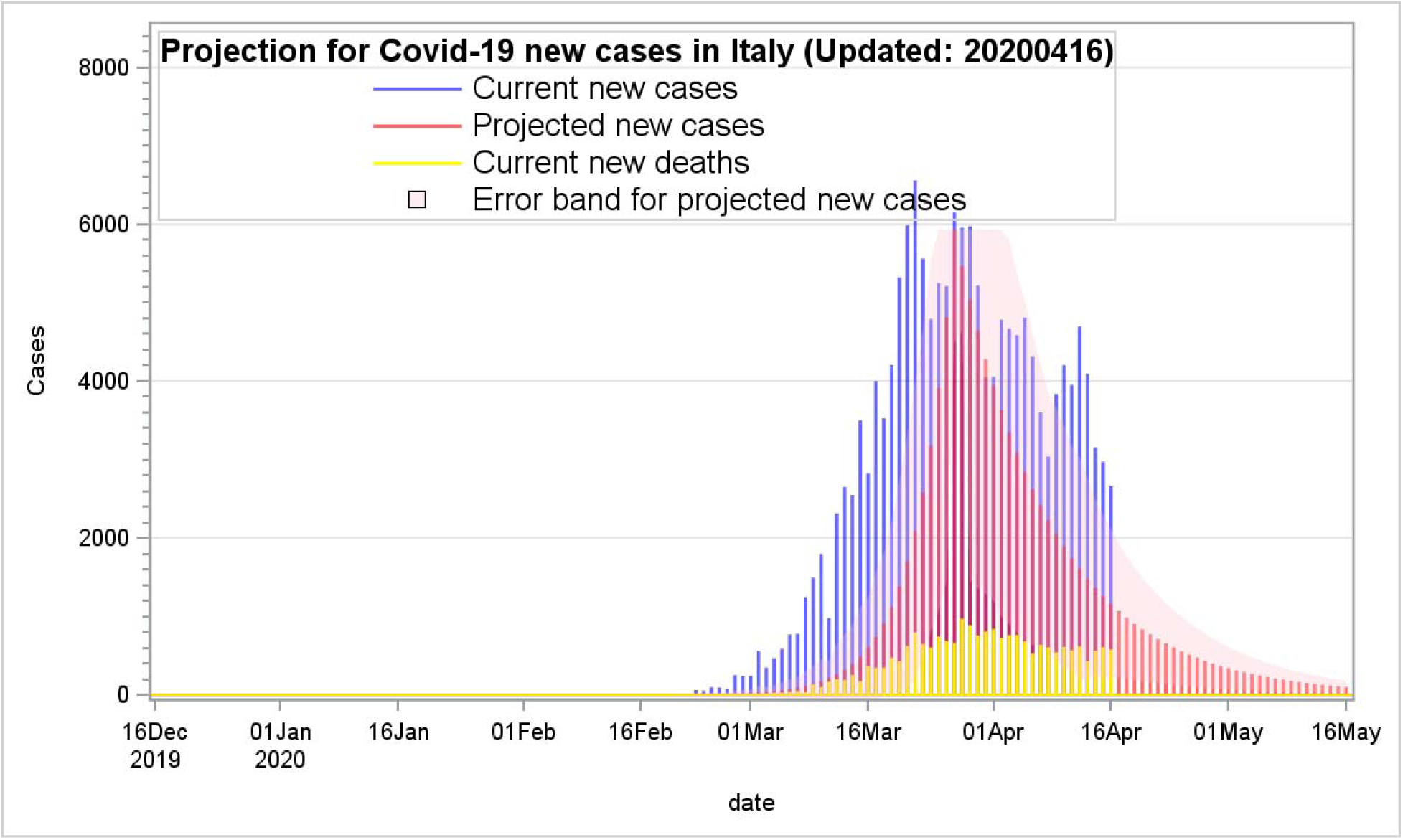
Projection for Covid-19 new cases in Italy.

**Figure 5.2:**
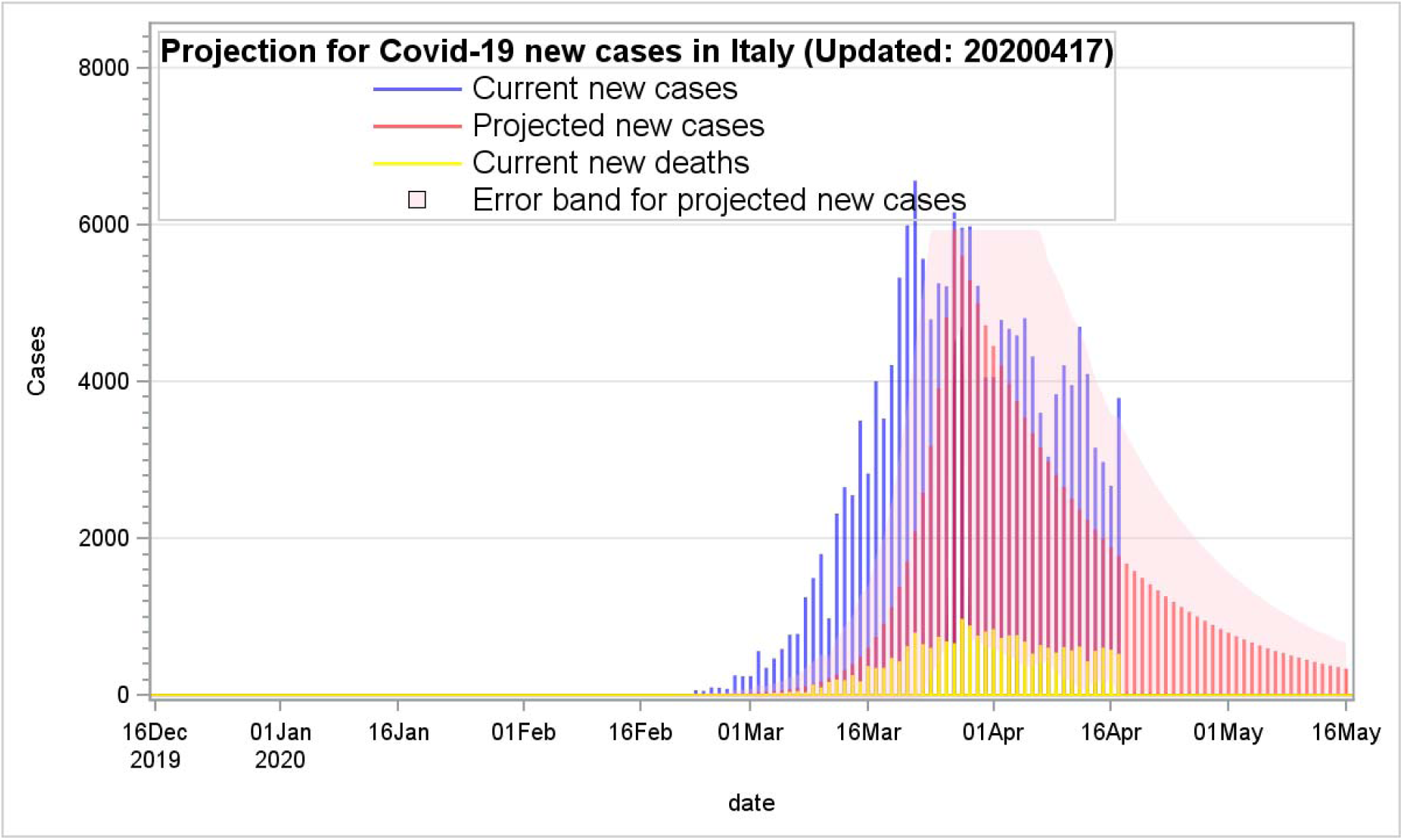
Projection for Covid-19 new cases in Italy.

### France

In France, new Covid-19 cases were beginning to increase dramatically at the end of February and then reaching to the apex of the outbreak up to 7,578 cases on April 1. The increasing trend seemed to follow along an exponential growth pattern. As shown in Figure 6.1, with the PPM estimates, the error band was accordingly expanded much higher after the much higher daily new 2,641 cases on April 17 were added into the PPM computations. As shown in Figure 6.2, the expanded error band with newly added data has improved the prediction precision for the upcoming days.

**Figure 6.1:**
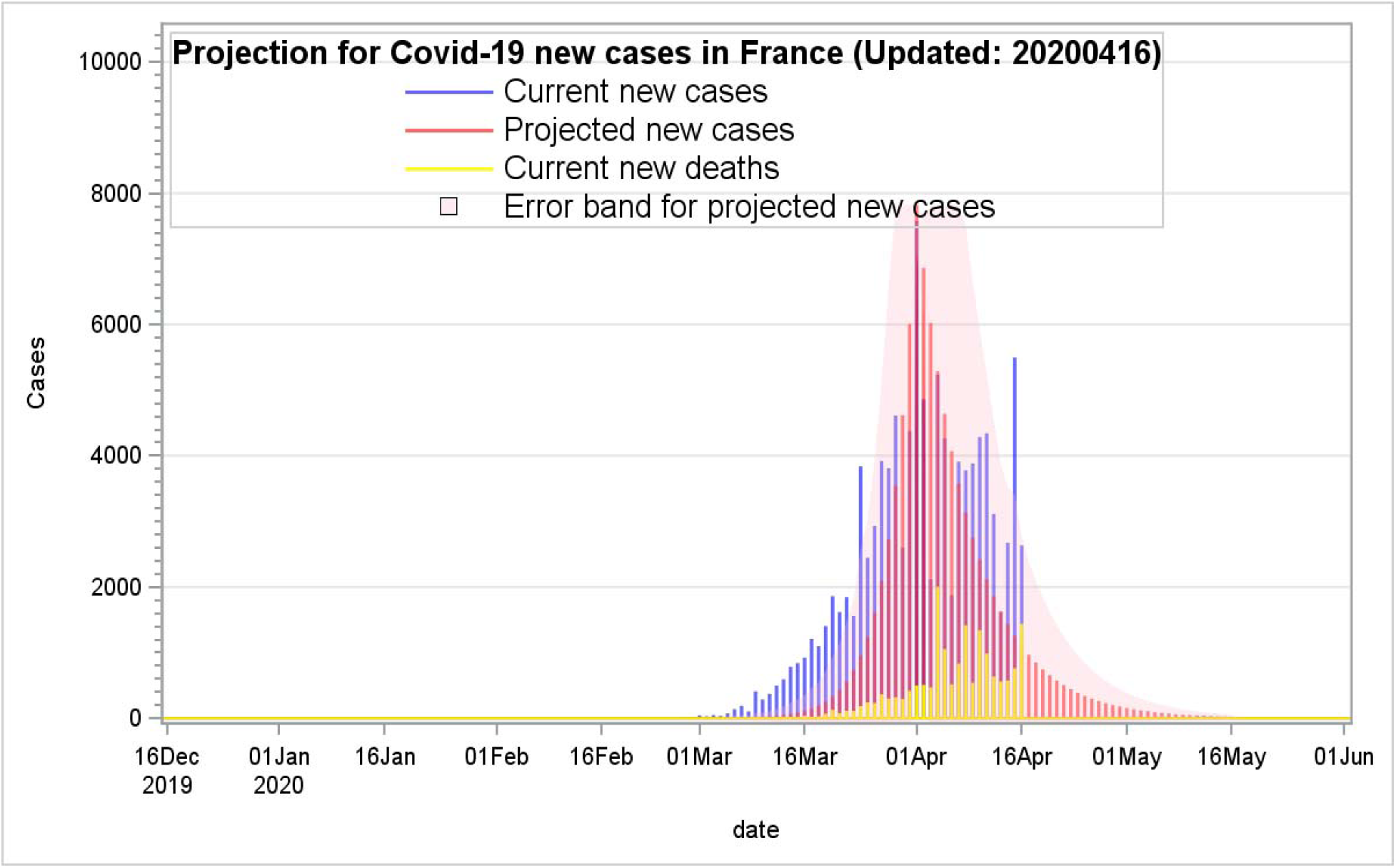
Projection for Covid-19 new cases in France.

**Figure 6.2:**
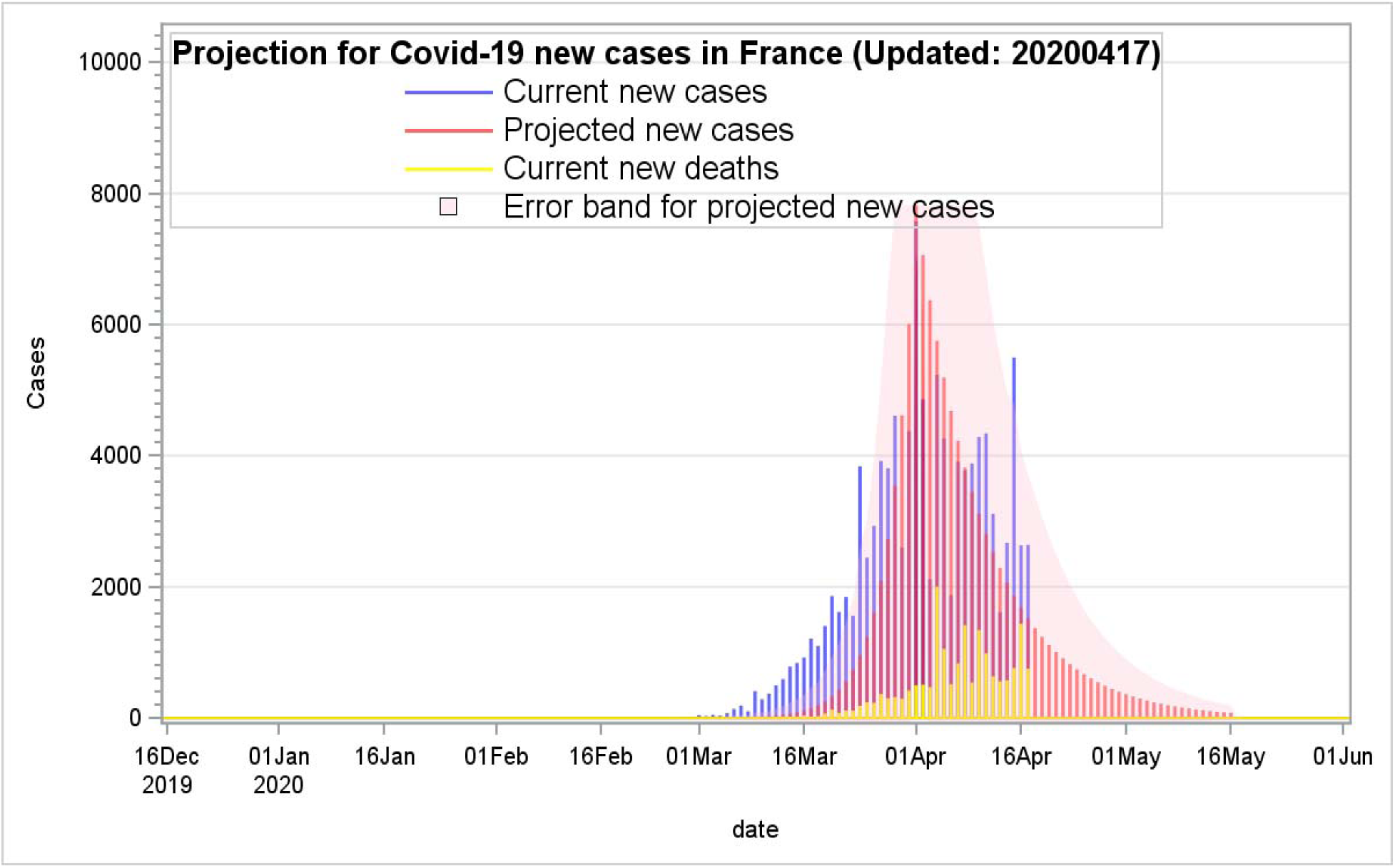
Projection for Covid-19 new cases in France.

### Strengths and limitations

I totally agree with Ferguson’s opinion on the role of mathematics modeling “models used for predictions are not crystal balls” [7]. The PPM is not primarily developed to target the peak day, rather it well predicts a plausible declining trend for the Covid-19 outlook. With the same concepts for the PPM computations, the numbers of new deaths, the needed hospital beds, and ventilations can be hopefully predictable if more data are collected from more regions or countries based on the city sizes, total population, population density, urbanization, by counting from the first onset day to the pandemic peak day. Furthermore, I agree with Bell’s comments “Many things can undermine a measurement. Measurements are never made under perfect conditions and in a laboratory, errors and uncertainties can come from [23]”. The development for the PPM is imperfect and not final and should be improved everlastingly. Especially, the improvement for the error band estimates is still very challenged as it is designed to have a better prediction with a wider error band boundary. However, but it might be over-inflated with some false alarms. Furthermore, to improve the prediction precisions, after passing the apex of the pandemic outbreak, multiple DHIs can be computed every 10 or more days to breakdown into more outbreak stages for improving the prediction precision.

## Discussions

The outbreak of Covid-19 has changed the human life forever. Its highly contagious and quick spread has a profound impact on how human work together, go shopping, and go to school without direct contact with each other. People are eager to know when the highly contagious disease will go away soon.

This study attempts to develop a plausible prediction method using the most current data. Based on the findings from the USA, four U.S. states (Illinois, Massachusetts, New Jersey, and New York), France, Italy, Spain, Germany, and China, the results indicated the PPM can provide quite accessible and reliable projections with the following reasons:

a. The PPM is adaptive and dynamic, because the simulations can be updated daily and renewed with new cases to improve its predictions.
b. This method uses live data for making more up-to-date predictions. Some models might only use hypothetical values and pure theoretical assumptions without using live data.
c. It is easy to understand and much accessible to replicate for multiple cities or states

On the other hand, the PPM computations should be kept updated daily. Without a frequent update, the prediction precision for the PPM will not be valid and reliable.

Many prevention methods by governments were used to control the widespread of Covid-19, such as the use of social distancing policy, face masks, self-quarantine, isolation, and bans for any travel from and to infected areas. Those factors should certainly have a great impact for the spread of the outbreak. The prediction models would not work well alone without considering many other factors together.

In the future, more web-based interactive platforms can be developed to quickly populate the PPM estimates for multiple big cities, regions, countries, or states to get more instant results for governments, health care professionals, or policymakers to help predict the outbreak trends earlier and to guide policymakers to prepare the worst ahead of time.

## Data Availability

The data used in this study are available for download

https://ourworldindata.org/coronavirus-source-data

https://github.com/nytimes/covid-19-data

## Funding

The author received no direct funding for this research.

**Compliance with ethical standards**

## Ethical approval

This study only used publicly available data and did not involve any experiments for human subjects.

## Informed consent

All data used were the numbers of confirmed Covid-19 cases without any human subject’s identify.

